# Medication adherence and hospitalizations in older patients with coronary heart disease in Vietnam

**DOI:** 10.1101/2024.09.23.24314196

**Authors:** Tan Van Nguyen, Nguyen Thi Thuy Hang, Truong Ngoc Dung, Nguyen Quoc Viet, Nguyen Quang Huy, Nguyen Quoc Huy, Ngo Thi Kim Trinh, Erkihun Amsalu, Wei Jin Wong, Tu Ngoc Nguyen

## Abstract

**Aim:** This study aimed to assess medication adherence among older people with coronary heart disease and its relationship with hospitalizations.

**Methods:** This is a prospective cohort study conducted at the outpatient clinics of a major hospital in Vietnam from November 2022 to June 2023. Consecutive older patients with coronary heart disease were recruited and followed for 6 months. Medication adherence was defined using the five[item Medication Adherence Report Scale (MARS-5). Multivariable logistic regression models were applied to examine the impact of medication adherence on hospitalization due to cardiovascular disease (CVD) and all-cause hospitalization.

**Results:** There were 643 participants. They had a mean age of 73 (SD 8), and 74.3% were male. Overall, 76.4% (491/643) were classified as “adherence”. Over 6 months follow-up, 23.3% of the participants admitted to hospital (9.2% were CVD-hospitalization). The CVD-hospitalization rate was significantly higher in the non-adherence group compared to the adherence group (13.8% versus 7.7%, p = 0.023, respectively). In logistic regression models, medication adherence was associated with a significant reduced odds of CVD hospitalization (adjusted OR 0.48, 95%CI 0.27 – 0.86). Medication adherence was also associated with a trend of reduced all-cause hospitalization (adjusted OR 0.75, 95%CI 0.49 – 1.15).

**Conclusions:** This study showed a positive relationship between medication adherence and reduced risk of CVD hospitalization in older people with coronary heart disease. Healthcare providers should consider incorporating adherence assessment into the long-term care for older patients with coronary heart disease.

## Introduction

Coronary heart disease is the leading cause of mortality globally.^1^ The prevalence of coronary heart disease increases with aging.^2^ The management of coronary heart disease aims to reduce cardiovascular events and mortality, and control symptoms and improve quality of life.^1^ The treatment of coronary heart disease is complex and generally includes a combination of optimizing risk factors, pharmacotherapy and percutaneous or surgical revascularisation if indicated. Pharmacological management includes anti-anginal medications (such as nitrates, beta blockers, calcium channel blockers), and cardioprotective medications (such as anti-platelets, statins, angiotensin-converting-enzyme inhibitors or angiotensin receptor blockers).^1^ These medications have been shown to reduce cardiovascular events in patients with coronary heart disease.^1^ In older people with coronary heart disease, medication adherence is critical to manage this condition. However, poor medication adherence continues to be a significant health challenge worldwide and many studies showed that medication adherence rates in older patients stand around 30% to 50%.^3,4^ Poor medication adherence can lead to worsening symptoms, increased risk of acute coronary syndromes and strokes, increased hospitalizations, and reduced overall quality of life.^5–8^ Poor or suboptimal medication adherence may also lead to unnecessary therapeutic intensification, whether dose increase or addition of new agents. This can further increase the risk of potential harm from polypharmacy. The risk is further amplified for older persons due to the biological changes associated with aging.

In Vietnam, the population is aging, and coronary heart disease is among the leading causes of mortality in this population.^9–12^ There have been several studies on medication adherence in patients with cardiovascular disease in Vietnamese adults. In a study of 175 patients (mean age 61) hospitalized due to acute myocardial infarction, the adherence rate to antiplatelet therapy among the participants was quite high at 1 month after discharge (90.3%), then declining by 6 months (88.0%), 12 months (75.4%), and more than 12 months (46.3%).^13^ A study conducted on 1038 patients with chronic cardiovascular disease (mean age 63) published in 2022 showed that only 59.3% of the participants were adherent to their cardiovascular medications.^14^ Another study in 177 patients (median age 63) on oral anticoagulants reported an adherence rate to oral anticoagulants of 37.7%.^15^ Study among people living in rural areas in Vietnam with hypertension (aged 35 to 64) showed that only 49.8% of the participants were adherent to antihypertensive medications.^16^ However, there is limited evidence on medication adherence in older people with coronary heart disease.

Therefore, this study aimed to assess adherence to cardiovascular medications in older people with coronary heart disease, and to examine the relationship between medication adherence and hospitalization in this population.

## Methods

### Study design and population

This prospective, observational study was conducted at the outpatient clinics of Thong Nhat Hospital in Ho Chi Minh City from November 2022 to June 2023. Consecutive patients aged ≥ 60 diagnosed with coronary heart disease who visited the clinics during the study period were recruited. Coronary heart disease was defined if a patient had any of these criteria in more than 3 months ago: (1) having a history of acute coronary syndrome, or (2) having significant stenosis on percutaneous coronary angiogram or computerized tomography coronary angiogram (≥ 50% for left main coronary artery, ≥ 70% for other coronary arteries), or (3) having percutaneous coronary interventions (PCI) or coronary artery bypass graft surgery (CABG). Exclusion criteria included: (1) having dementia or having a mental illness that can affect their ability to answer the study questionnaires, (2) not being able to provide consent, and (3) having a life expectancy <6 months.

The study was approved by the Ethics Committees of the University of Medicine and Pharmacy at Ho Chi Minh City (Reference Number 936/HDDD-DHYD, date 24/11/2022). Informed consent was obtained from all participants. This study was compiled in accordance with the Declaration of Helsinki.

### Data collection

Data were collected from patient interviews and medical records. Information obtained included demographic characteristics, height, weight, medical history, blood test results, and comorbidities. Frailty was assessed using the Clinical Frail Scale (CFS).^17,18^ The CFS score ranges from 1-9, and a score of 4 or greater indicates a frailty status.^17,19^ Polypharmacy was defined as using 5 or more medications on a daily basis. Cardiovascular multimorbidity (CVD multimorbidity) was defined as having any of the following conditions in addition to coronary heart disease: heart failure, stroke, atrial fibrillation, peripheral artery disease, chronic kidney disease, and diabetes.

Assessment of medication adherence: Participants’ medication adherence was assessed using the five[item Medication Adherence Report Scale, MARS-5 (© Professor Rob Horne).^20,21^ We use the Vietnamese version of the MARS-5 which was approved by Professor Rob Horne. The MARS-5 questionnaire comprised five components: (1) I forget to take my medicines, (2) I alter the dose, (3) I stop taking them for a while, (4) I decide to miss out a dose, (5) I take less than instructed. The first statement indicates unintentional non-adherence, and the other four statements indicate intentional non-adherence. The participants answered these five statements on a 5-point Likert scale (1 = always, 2 = often, 3 = sometimes, 4 = rarely, 5 = never). MARS-5 total scores range from 5 to 25, and higher scores indicate better medication adherence. In line with previous studies, we use a cut-off value of 23 to define adherence: non-adherence was defined as MARS-5 scores ≤ 23 and adherence was defined as MARS-5 scores 24–25.^20,21^

Outcome variables: The primary outcome was CVD hospitalization. The secondary outcome was all-cause hospitalization. All participants were followed up for 6 months after being included in the study. Hospitalization information was obtained by making phone calls to the phone numbers provided by participants or their caregivers after 6 months. The causes of hospitalization were documented and classified as cardiovascular disease related hospitalization (CVD hospitalization) or all-cause hospitalization.

### Sample size estimation

Based on the local data, we estimated that the rate of CVD hospitalization in older patients with chronic coronary heart disease in 6 months would be around 10%. Therefore, we estimated that at least 640 patients with coronary heart disease would be needed in this study to detect a difference in the CVD hospitalization rates between patients who were adherent to medication compared to those who were non-adherent (assuming a relative difference of 40% in CVD hospitalization rates between the two groups, with a power of 80%, 1-sided test, alpha=0.05, and allowing for 10-12% drop out during follow up).

### Statistical analysis

Study population characteristics are presented as mean and standard deviation (SD) for continuous variables, or frequencies and percentages for categorical variables. Comparisons in general characteristics and hospitalization rates between the adherence and non-adherence groups were conducted using Chi-square tests or Fisher’s exact test for categorical variables, and Student’s t-tests for continuous variables. Multivariable logistic regression models were applied to examine the impact of medication adherence on CVD hospitalization and all-cause hospitalization. The following covariates were hypothesized to possibly have an impact on hospitalization in older people with coronary heart disease and were therefore included in the adjusted logistic regression models: age, sex, history of PCI/CABG, frailty, and CVD multimorbidity. Age (in years) and frailty (the CFS score) were treated as continuous variables, and all other variables were categorical. P values <0.05 were considered statistically significant. Data were analyzed in IBM SPSS Statistics 27.0. and R 4.3.1.

## Results

A total of 643 participants were included in this study. They had a mean age of 73.1, 25.7% were female and 74.3% were male. **Table 1** presents the participant characteristics. A majority of participants (49.0%) had completed higher education. Most of the participants retired, and 4.4% of them were still working. The mean CFS score was 3.9, and 60.3% of the participants were classified as being frail. Polypharmacy was presented in 89.6% of the participants (with a mean total number of medications = 6.7), and 65.8% had a history of PCI/CABG. Hypertension and dyslipidemia were present in 96.9% and 95.3% of the participants, respectively. Among the cardio-metabolic comorbidities, diabetes was the most prevalent (42.6%), followed by atrial fibrillation (30.1%), heart failure (15.6%), chronic kidney disease (8.0%), and ischemic stroke (5.6%). Regarding carer, 10.6% of the participants did not have any carer, 70.6% had support from their spouse, 17.0% from their children, and 1.8% from other relatives or professional carers.

**Table 1.** Participant characteristics.

| Characteristics | All participants<br>(N = 643) | Non-adherence<br>group<br>(N = 152) | Adherence<br>group<br>(N = 491) | P |
| --- | --- | --- | --- | --- |
| Age | 73.1 ± 8.3 | 72.0 ± 7.7 | 73.5 ± 8.4 | 0.064 |
| Age group |  |  |  | 0.104 |
| 60 – 69 | 255 (39.6%) | 71 (46.7%) | 184 (37.5%) |  |
| 70 – 79 | 237 (36.9%) | 52 (34.2%) | 185 (37.7%) |  |
| ≥ 80 | 151 (23.5%) | 29 (19.1%) | 122 (24.8%) |  |
| Sex |  |  |  | 0.011 |
| Female | 165 (25.7%) | 27 (17.8%) | 138 (28.1%) |  |
| Male | 478 (74.3%) | 125 (82.2%) | 353 (71.9%) |  |
| Working status |  |  |  | 0.530 |
| Retired | 615 (95.6%) | 144 (94.7%) | 471 (95.9%) |  |
| Working | 28 (4.4%) | 8 (5.3%) | 20 (4.1%) |  |
| Carer |  |  |  | 0.006 |
| None | 68 (10.6%) | 20 (13.2%) | 48 (9.8%) |  |
| Spouse | 454 (70.6%) | 118 (77.6%) | 336 (68.4%) |  |
| Children | 109 (17.0%) | 12 (7.9%) | 97 (19.8%) |  |
| Other | 12 (1.8%) | 2 (1.3%) | 10 (2.0%) |  |
| Public health insurance |  |  |  | 0.667 |
| Yes | 632 (98.3%) | 150 (98.7%) | 482 (98.2%) |  |
| No | 11 (1.7%) | 2 (1.3%) | 9 (1.8%) |  |
| Education (missing 31) |  |  |  | 0.184 |
| Illiterate | 16 (2.5%) | 7 (4.7%) | 9 (1.9%) |  |
| Primary school/Secondary school | 110 (18.1%) | 22 (14.8%) | 88 (19.0%) |  |
| High school | 186 (30.4%) | 43 (28.9%) | 143 (30.9%) |  |
| Higher education | 300 (49.0%) | 77 (51.7%) | 223 (48.2%) |  |
| Body mass index |  |  |  | 0.291 |
| Underweight | 36 (5.6%) | 7 (4.6%) | 29 (5.9%) |  |
| Normal | 223 (34.7%) | 46 (30.3%) | 177 (36.0%) |  |
| Overweight | 189 (29.4%) | 44 (28.9%) | 145 (29.5%) |  |
| Obese | 195 (30.3%) | 55 (36.2%) | 140 (28.5%) |  |
| Smoking |  |  |  | 0.066 |
| Non-smoking | 327 (50.9%) | 67 (44.1%) | 260 (53.0%) |  |
| Current smoking | 70 (10.9%) | 23 (15.1%) | 47 (9.6%) |  |
| Ex-smoking | 246 (32.2%) | 62 (40.8%) | 184 (37.5%) |  |
| Frailty (CFS $\geq 4$) | 388 (60.3%) | 92 (60.5%) | 296 (60.3%) | 0.958 |
| CFS score | 3.9 ± 1.3 | 3.8 ± 1.2 | 3.9 ± 1.3 | 0.721 |
| Total number of medications | 6.7 ± 1.8 | 7.2 ± 4.7 | 6.7 ± 3.0 | 0.132 |
| Polypharmacy (using $\geq 5$ medications) | 576 (89.6%) | 141 (92.8%) | 435 (88.6%) | 0.142 |
| Medical history: |  |  |  |  |
| PCI | 413 (64.3%) | 102 (67.1%) | 311 (63.3%) | 0.397 |
| CABG | 10 (1.5%) | 2 (1.3%) | 8 (1.6%) | 0.785 |
| Hypertension | 623 (96.9%) | 150 (98.7%) | 473 (96.3%) | 0.145 |
| Dyslipidemia | 613 (95.3%) | 144 (94.7%) | 469 (95.5%) | 0.689 |
| Diabetes | 274 (42.6%) | 58 (38.2%) | 216 (44.0%) | 0.204 |
| Atrial fibrillation | 195 (30.1%) | 34 (22.4%) | 161 (32.8%) | 0.015 |
| Peripheral artery disease | 164 (25.7%) | 45 (29.8%) | 119 (24.4%) | 0.183 |
| Heart failure | 100 (15.6%) | 26 (17.1%) | 74 (15.1%) | 0.545 |
| Chronic kidney disease | 52 (8.0%) | 11 (7.2%) | 41 (8.4%) | 0.660 |
| Ischemic stroke | 36 (5.6%) | 9 (5.9%) | 27 (5.5%) | 0.843 |
| Cardiovascular multimorbidity | 511 (79.5%) | 113 (74.3%) | 398 (81.1%) | 0.073 |

Continuous data are presented as mean ± standard deviation. Categorical data are shown as n (%). PCI, percutaneous coronary intervention; CABG, coronary artery bypass graft surgery; CFS, Clinical Frailty Scale.

Compared to the non-adherence group, the adherence group was older (mean age 73.5 in the adherence group vs. 72.0 in the non-adherence group). The proportion of females was higher in the adherence group (28.1% vs. 17.8% in the non-adherence group, p = 0.011). The proportion of having support from children was significantly higher in the adherence group compared to the non-adherence group (19.8% vs. 7.9%, p = 0.006, respectively). There was no significant difference in the prevalence of cardiovascular risk factors and cardiovascular comorbidities between the two groups, except for atrial fibrillation, which was more prevalent in the adherence group (32.8% vs. 22.4% in the non-adherence group, p = 0.015)

### Medication adherence

The distribution of the MARS-5 scores and the individual components are presented in **Table 2**. Overall, 76.4% (491/643) were classified into the adherence group, and 23.6% (152/643) of the participants were classified into the non-adherence group.

**Table 2.** Distribution of the MARS-5 scores and its individual components.

| | | Always<br>= 1<br>(n, %) | Often<br>= 2<br>(n, %) | Sometimes<br>= 3<br>(n, %) | Rarely<br>= 4<br>(n, %) | Never<br>= 5<br>(n, %) | Mean ± SD |
| --- | --- | --- | --- | --- | --- | --- | --- |
| Item 1 | I forget to take my medicines | 0 | 12 (1.9%) | 117 (18.2%) | 37 (5.8%) | 477 (74.2%) | 4.52 ± 0.85 |
| Item 2 | I alter the dose | 0 | 3 (0.5%) | 9 (1.4%) | 6 (0.9%) | 625 (97.2%) | 4.95 ± 0.32 |
| Item 3 | I stop taking them for a while | 0 | 4 (0.6%) | 8 (1.2%) | 6 (0.9%) | 625 (97.2%) | 4.95 ± 0.34 |
| Item 4 | I decide to miss out a dose | 0 | 5 (0.8%) | 10 (1.6%) | 8 (1.2%) | 620 (96.4%) | 4.93 ± 0.38 |
| Item 5 | I take less than instructed | 0 | 4 (0.6%) | 15 (2.3%) | 15 (2.3%) | 609 (94.7%) | 4.91 ± 0.41 |

### Hospitalization

Over 6 months of follow-up, 23.3% of the participants were admitted to hospitals (22.4% in the adherence group compared to 26.3% in the non-adherence group, p=0.319). Of these, 9.2% were due to CVD. The CVD-hospitalization rate was significantly higher, almost doubled, in the non-adherence group compared to the adherence group (13.8% versus 7.7%, p = 0.023, respectively) (**Figure 1**)

**Figure 1.**
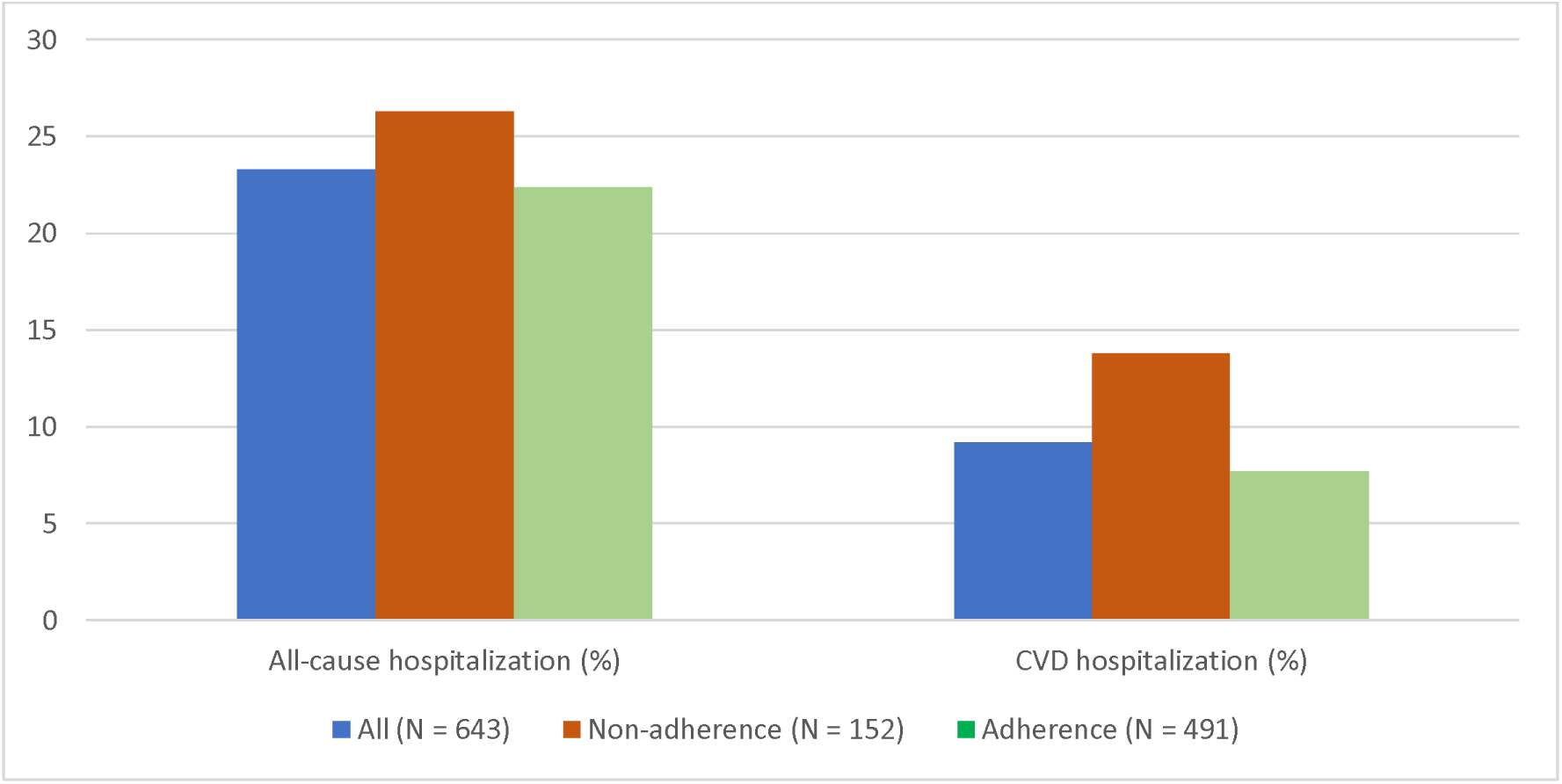
Hospitalization rates after 6 months.

### The relationship between adherence and hospitalization

In logistic models, medication adherence was associated with significantly reduced CVD hospitalization likelihood (adjusted OR 0.48, 95%CI 0.27 – 0.86). A history of PCI/CABG was also associated with reduced CVD hospitalization (adjusted OR 0.53, 95%CI 0.29-0.94). (**Figure 2**)

**Figure 2.**
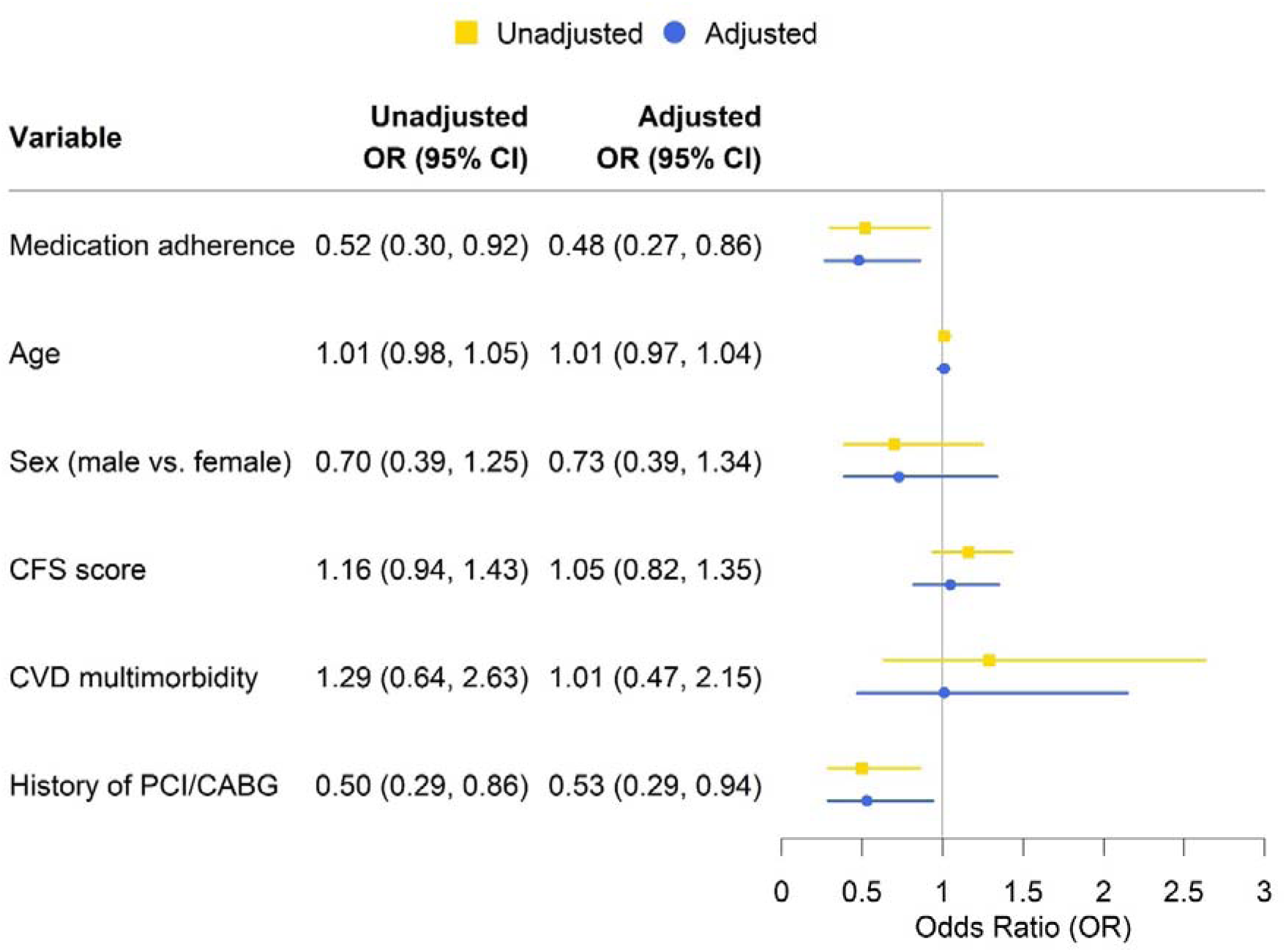
Predictor factors for CVD hospitalization. CFS: Clinical Frailty Scale. CVD: cardiovascular disease. PCI: percutaneous coronary interventions. CABG: coronary artery bypass graft surgery.

Medication adherence was also associated with a trend of reduced all-cause hospitalization, but the difference was not statistically significant (adjusted OR 0.75, 95%CI 0.49 – 1.15). In the adjusted model, frailty was the only independent predictor for all-cause hospitalization (adjusted OR 1.19, 95%CI 1.01-1.41 for every 1-point increase in the CFS score). (**Figure 3**)

**Figure 3.**
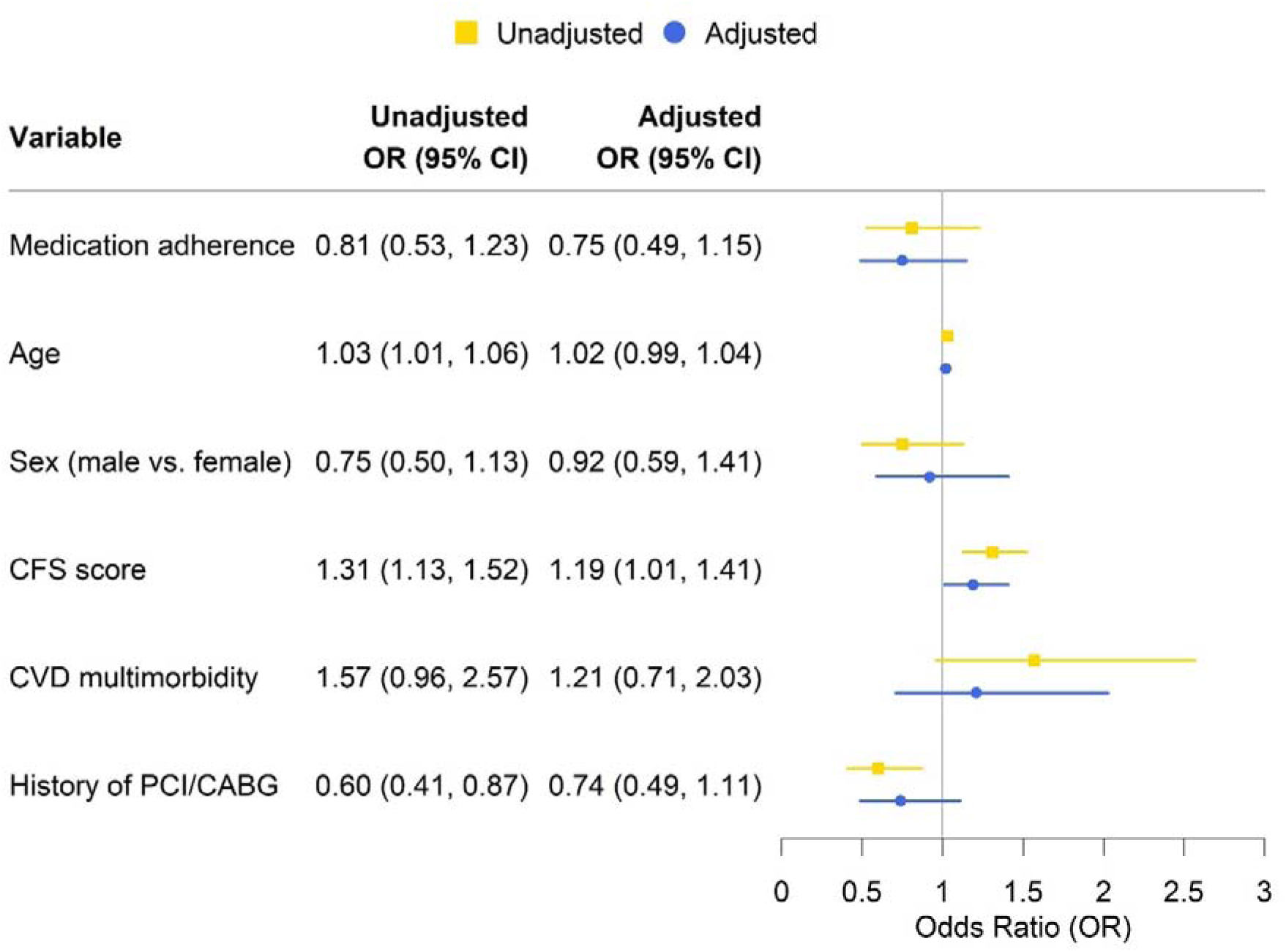
Predictor factors for all-cause hospitalization. CFS: Clinical Frailty Scale. CVD: cardiovascular disease. PCI: percutaneous coronary interventions. CABG: coronary artery bypass graft surgery.

## Discussion

Our study evaluated medication adherence in older people with coronary heart disease. The study contributed to the limited existing evidence on medication adherence in Vietnamese people by providing insights for the older population. Our analysis showed that a high proportion of older participants with chronic coronary heart disease in the study reported being adherent to their medications. We found an association between medication adherence and reduced hospitalization rates, particularly CVD related hospitalization.

Information on medication adherence is important as it contributes to patient health. Published studies on medication adherence in the Vietnamese population showed varying levels of adherence, depending on the patient cohorts, clinical settings and the instruments used to measure and define adherence.^13–16^ For example, a study using the General Medication Adherence Scale (GMAS) reported a rate of 40.7% of non-adherence in Vietnamese patients with chronic cardiovascular diseases.^14^ This rate is higher compared to our study, and this could be due to the mean age of that study population being younger compared to our study (62.7 years of age vs 73.1 years of age in our study).^14^ Better medication adherence with increasing age has been reported in a previous study in Vietnam.^16^ While most previous studies on medication adherence in Vietnam recruited younger participants (age ranged from 53 to 63),^13–16^ the result of our study provides insights on medication adherence for the older population. Vietnam is going through an epidemiological transition with a growing number of older people. The older population typically requires increased utilization of healthcare resources due to their more significant health needs. Therefore, the findings of our study can provide valuable guidance for developing health policies and plans at a population level, particularly in settings where resources are limited.

Studies in neighbouring low to middle-income countries with similar health systems and demographic challenges like Vietnams also report varying levels of adherence and implications. For example, a study in a primary care clinic in Malaysia reported fairly good adherence (60.3%) in patients with type 2 diabetes mellitus (mean age 60 years), although chronological age was significantly associated with poorer medication adherence.^22^ Patients with different comorbidities may have different health goals and challenges which could influence their perception of medication adherence. In this case, patients with type 2 diabetes may have different perspectives towards their medications when compared to patients with coronary heart disease. As patients with diabetes grow older, their perceptions towards the risk of adverse effects from their medications may influence their adherence levels. Additionally, patients with type 2 diabetes sometimes practice self-adjustment of dosage regimes, which may then also affect adherence levels. In another study of Thai patients with diabetic chronic kidney disease (mean age 71 years), approximately 50% of the study participants were classified as having medium to low medication adherence, and low medication adherence was associated with poorer blood pressure control and poorer kidney outcome.^23^ This further highlights the influence of medication adherence on health outcomes and the different experiences of patients with different chronic health conditions. With the growing older population in these countries and surrounding areas, further studies are needed to help guide the development and provision of targeted care in the different sub-group populations.

Our study results showed that medication adherence was associated with reduced hospitalization rates, particularly CVD hospitalization. The risk of CVD hospitalization was reduced by half in the adherence group compared to the non-adherence group in our study. This finding is in line with observations from studies worldwide. In a systematic review on the impact of medication adherence on coronary heart disease costs and outcomes conducted by Bitton and colleagues, the authors found that high adherence was significantly associated with reduced coronary artery disease-related events, mortality, readmissions, and annual costs for secondary prevention of coronary artery disease.^24^ Cardiovascular diseases continue to be a significant source of burden of disease on health systems in low to middle-income countries like Vietnam.^25^ The growing impact of CVD on health systems shows the need for more steps to be taken to help manage this. Pharmacotherapies continue to play essential roles in managing coronary heart disease in older people and as such, medication adherence should be an integral part of broader management strategies. Importantly, medication adherence is a modifiable behavioural risk factor that can be targeted for intervention. There is a pressing need for innovative and evidence-based strategies to enhance medication adherence in older patients with coronary heart disease in Vietnam. Medication non-adherence is pervasive globally and more so in low- and middle-income countries that have fewer healthcare resources.^26^ Association of medication adherence with major adverse cardiovascular events also have implications for healthcare costs, which is a major challenge for policymakers in resource limited settings. As the management of CVD involves high utilization of healthcare resources, focusing on medication adherence as a modifiable patient factor becomes a possible focus point when planning cost-effective strategies for managing cardiovascular disease.

Exploring factors for non-adherence could also potentially reduce unnecessary hospital readmission rates. Considering the broader level impact of medication adherence, more studies should be done to explore factors influencing medication adherence in Vietnam, particularly in the older population. Older patients with coronary heart disease are more likely to experience multiple chronic conditions and polypharmacy, making it challenging to adhere to complex medication regimens. In addition, physiological changes associated with aging, such as cognitive decline and physical limitations, can also impact medication adherence in older individuals, especially those living in low- and middle-income countries.^27,28^ Patients in low and middle-income countries have added barriers compared to patients in high-income or developed countries. Examples of these barriers may include limited access to medicines, varying educational backgrounds affecting health literacy levels, and resources to support medication adherence. A recent systematic review of factors that can influence medication adherence of adults with chronic diseases found that socioeconomic status and social support might have a positive impact on adherence.^29^ In patients taking long-term medicines for CVD prevention, their perception of health goals may affect medication adherence.^30^ In low- and middle-income countries, poor medication adherence has also been influenced by a lack of knowledge, negative beliefs, and negative attitudes.^28^ As young and old patients may have different perceptions of health goals, further studies could be done to explore specific reasons for non-adherence in older patients with coronary heart disease. Further studies could also be conducted to explore the relationship of educational background, its impact on health literacy and its relationship with factors experienced by the older population such as frailty and polypharmacy.

To the best of our knowledge, this was the first study to examine medication adherence in older people with chronic coronary syndrome in Vietnam. Additionally, our study was conducted at Thong Nhat Hospital, one of the largest hospitals in Vietnam with specialized services for older patients, which enabled the collection of data for older people with complex medication management plans. Our study also reported the relationship with hospitalization rates, an outcome that is not routinely reported in studies on medication adherence. With the growing population of older people coupled with the growing burden of cardiovascular diseases, factors that can influence hospitalization rates should be considered for healthcare planning. This becomes more important in health systems with limited health resources where there is added pressure of juggling the multiple health challenges it faces.

However, our study had several limitations. The study was conducted on older patients attending outpatient clinics from one hospital, so it may not accurately reflect all older people with coronary heart disease in Vietnam. There was a high proportion of achieving higher education among our study participants, which may explain the high proportion of medication adherence observed in this study. In addition, our follow-up duration was only 6 months, and we did not re-assess medication adherence at the end of the follow up. Further studies with larger sample sizes and longer follow-ups are needed to understand the impact of medication adherence on adverse outcomes and quality of life in older patients with coronary heart disease.

## Conclusion

This study showed a positive relationship between medication adherence and reduced risk of CVD hospitalization in older people with coronary heart disease. Our study highlights the important role of medication adherence in improving health outcomes for older people with coronary heart disease. There is a need to better understand the reasons why patients are non-adherent to their medicines, particularly in different contexts and specific populations. Contextualized strategies would be useful to help improve adherence and potentially reduce unnecessary hospitalization rates. Healthcare providers should consider incorporating adherence assessment into the long-term care for older patients with coronary heart disease.

## Data Availability

All data produced in the present study are available upon reasonable request to the authors.

## Acknowledgement

There was no funding provided for this study.

## Author Contribution

T.V.N. is the Principal Investigator and oversaw the study. T.V.N and T.N.N. led the study concept and study design, and wrote the first draft of the manuscript. T.V.N., N.T.T.H, T.N.D, N.Q.V., N.Q.H., N.Q.H led ethics application, recruitment, and data acquisition. All authors (T.V.N., N.T.T.H, T.N.D, N.Q.V., N.Q.H., N.Q.H, N.T.K.T., E.A., W.J.W., T.N.N.) were involved in analysis and interpretation of data, and revised the manuscript critically for important intellectual content. All authors read and approved the final manuscript.

## Data availability statement

The data that support the findings of this study are available on request from the corresponding author. The data are not publicly available due to privacy or ethical restrictions.

